# Radiological follow-up of adults hospitalised with pneumonia and SARS-CoV-2 infection, in Bristol UK, during the COVID19 pandemic

**DOI:** 10.1101/2022.01.04.22268738

**Authors:** Gabriella Ruffino, Rachel L Williams, Shaney Barratt, Catherine Hyams

**Author notes:** Corresponding Author: Dr Catherine Hyams, MBBS BSc(Hons) PhD, Academic Respiratory Unit, Learning and Research, Southmead Hospital, Bristol, UK, BS10 5NB. **Funding:** CH was funded by the National Institute for Health Research [NIHR Academic Clinical Fellowship (ACF-2015-25-002). The views expressed are those of the author(s) and not necessarily those of the NIHR or the Department of Health and Social Care.

## Abstract

**Introduction:** Radiological change which may be attributed to infection can also be attributable to lung cancer. Patients with SARS-CoV-2 infection can develop groundglass lung opacification which may result in chronic lung changes. Current British Thoracic Society (BTS) guidelines recommend patients with pneumonia and COVID19 undergo repeat chest radiography.

**Methods:** A single-centre audit of patients hospitalised with community-acquired pneumonia or COVID19 over three time periods during the COVID19 pandemic (Aug-Dec 2020, Jun-Aug 2021, Dec-Jan 2022). We assessed whether patients were eligible for radiological follow-up and if repeat radiological investigation occurred.

**Results:** 1040 adults were hospitalised with infective radiological change (pneumonia=596, COVID19=444). 831/1040 patients (80%) required radiological follow-up under BTS guideline criteria: there was minimal difference between the first two time periods studied. Patients hospitalised with CAP were less likely to have radiological follow-up planned than those admitted with COVID19 disease (49% versus 59% respectively). Following a change in hospital policy, follow-up rates increased to 69% and 71% for pneumonia and COVID19. Overall, only 47% eligible patients received follow-up in line with current guidelines.

**Conclusion:** BTS guideline adherence is important to avoid delay in diagnosing underlying malignancy or chronic lung disease. Radiological follow-up following CAP and COVID19 may be suboptimal, with a paucity of data. Follow-up arranged under the hospital team was more likely to occur than when the GP was responsible for instigating repeat radiological imaging. Further investigation into rates of radiological follow-up should be undertaken, including reasons for non-adherence, to ensure patients receive appropriate treatment following respiratory infection.

## INTRODUCTION

Pneumonia causes significant morbidity and mortality, and even before the COVID19 pandemic the healthcare cost of pneumonia in Europe was estimated at approximately €10 billion per annum, with inpatient care accounting for €5.7 billion^1^. Community-acquired pneumonia (CAP) has an incidence of 7.99 per 1,000 adults in the UK with disease dramatically increasing with age, particularly in >85-year-olds^2^. However, radiological change which may be attributed to infection (Figure 1A) can also be attributable to lung cancer, and obstructing lung lesions are an underlying cause of pneumonia. Furthermore, obstructing lung lesions can cause pneumonia, and lung cancer remains a leading cause of cancer-related death^3^. Notably, 2.5% of chest radiographs repeated <90 days of pneumonia diagnosis result in a new diagnosis of lung cancer, with cancer diagnostic predicted to increase if radiographs were only repeated in patients >50 years^4^. Therefore British Thoracic Society (BTS) guidelines recommend repeating chest radiography following CAP diagnosis at 6-weeks in patients ‘at higher risk of underlying malignancy’^5^.

**Figure 1:**
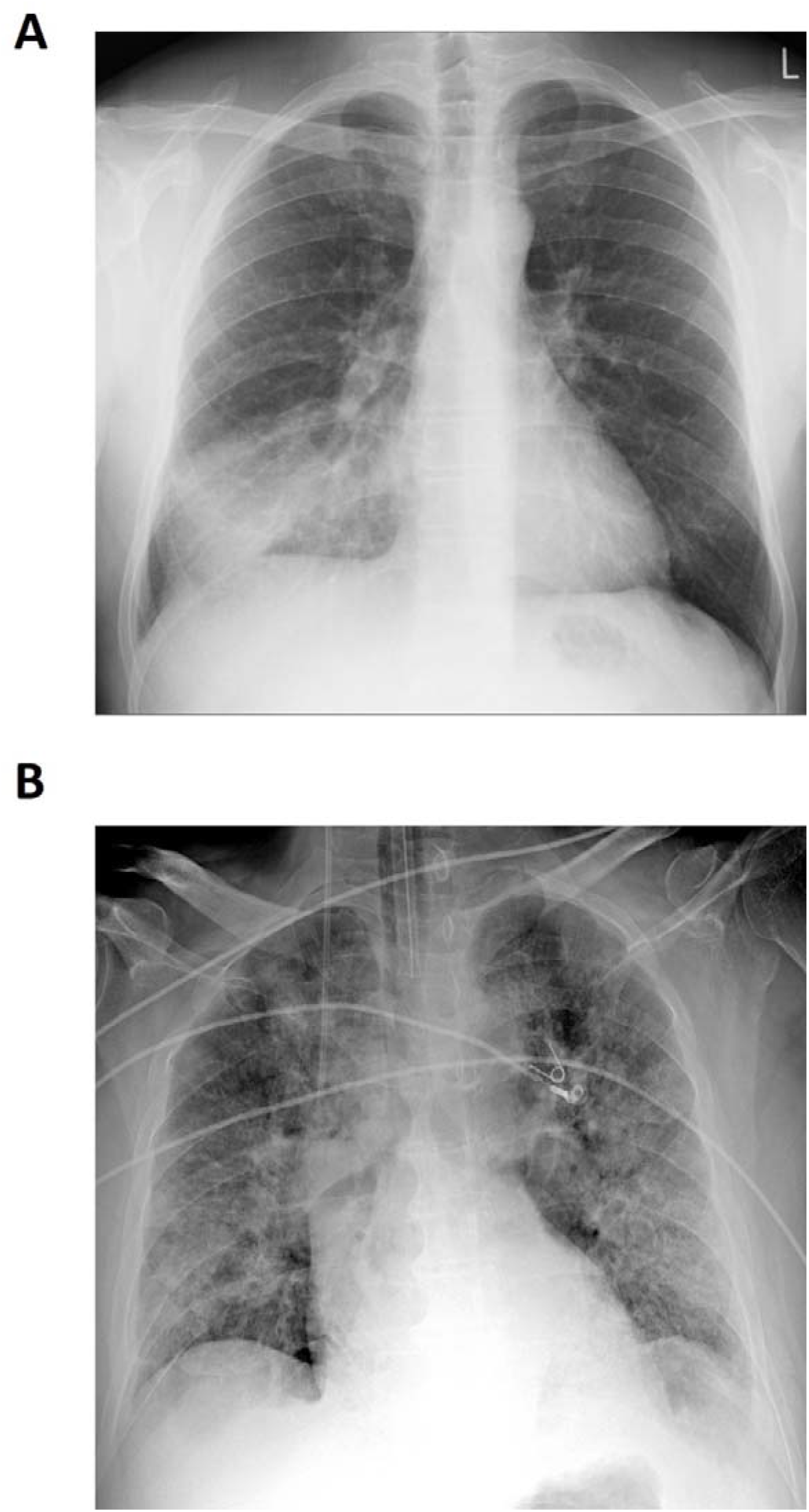
Radiological features of pneumonia and SARS-CoV-2 infection. (A) AP chest radiograph showing right middle lobe consolidation (25) and (B) extensive bilateral ground-glass opacities in a patient with confirmed SARS-CoV-2 infection (26).

Severe acute respiratory syndrome Coronavirus 2 (SARS-CoV-2) has emerged as a new and highly contagious respiratory pathogen, causing COVID19. Patients with COVID19 are reported to exhibit characteristic radiological changes of groundglass lung opacification (Figure 1B), potentially associated with linear opacities, and which may progress to bilateral consolidation^6^. These radiological changes following SARS-CoV-2 infection are similar to those occurring with other coronaviruses, which can result in chronic lung changes on imaging and reduced lung function^7,8^. Recent BTS guidelines recommend repeating radiology at 12-weeks post-discharge for patients with a COVID19 diagnosis^9^. We therefore audited patients hospitalised with lobar consolidation due to CAP and opacities consistent with COVID19 pneumonitis to determine current BTS guideline adherence.

## METHODS

Adults ≥18 years hospitalised to North Bristol NHS Trust were audited (1) from 1st August until 30^th^ November 2020, (2) 1^st^ June until 31^st^ July 2021 and (3) 28^th^ Dec to 19^th^ Jan 2022. These windows reflect time periods during which hospitalisations with COVID19 were increasing in Bristol (Supplementary Data 1) due to the emergence of Alpha or Delta variants, and in a period of established Omicron dominance (Figure 2). The experience of clinical staff and familiarity with treatment protocols would have increased between period 1 and 2, as doctors rotate in August in the UK. Between the second and third audit period, the radiological follow-up of patients with CAP or COVID19 became the responsibility of the treating inpatient medical team.

**Figure 2:**
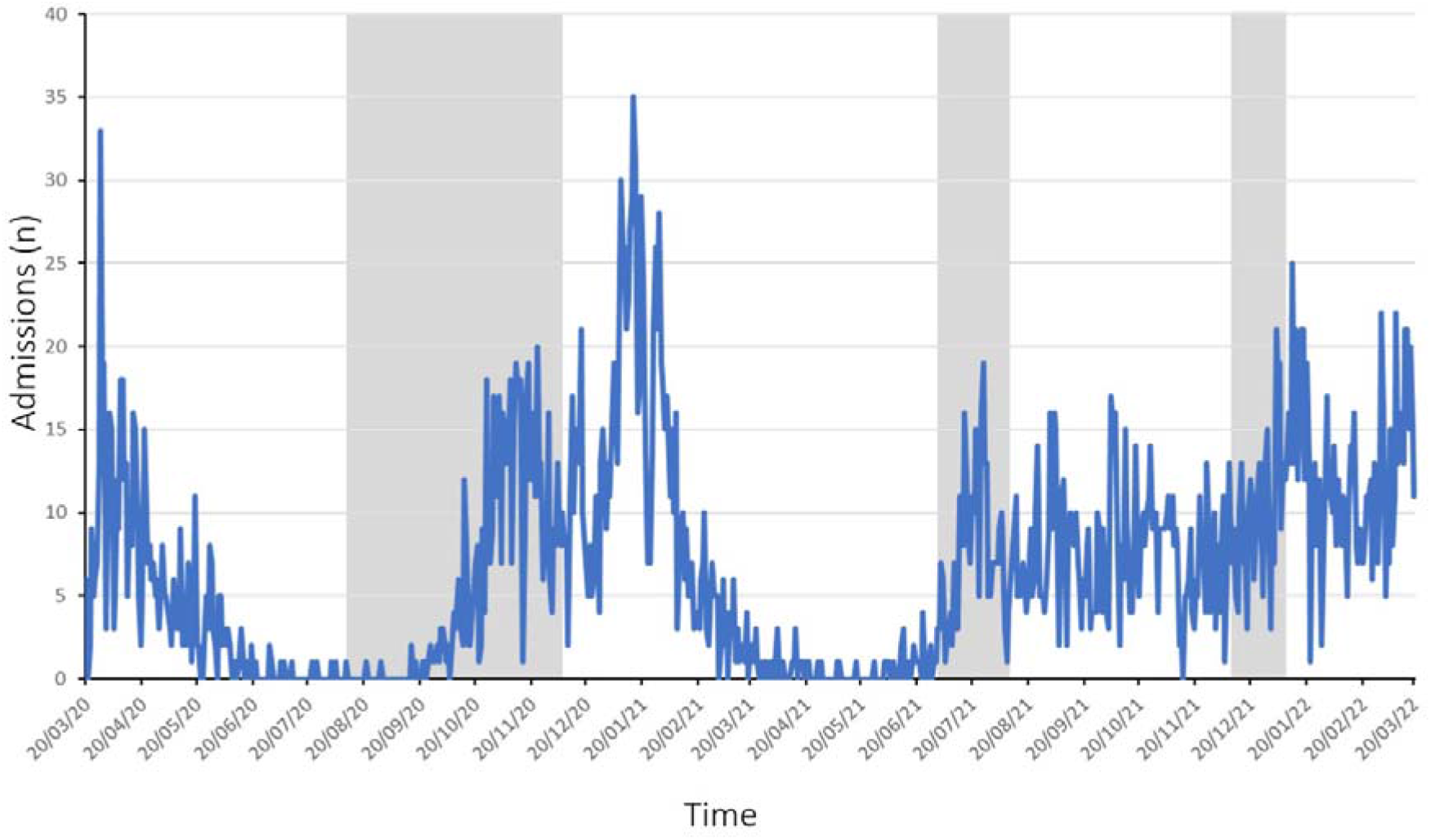
Hospital admissions with SARS-CoV-2 infection. Daily admissions to North Bristol NHS Trust with SARS-CoV-2 infection are shown (blue line) from 20 March 2020 until 20 March 2022. The grey bars represent the periods of time during which this audit was undertaken at the hospital. Data was obtained from UK COVID19 dashboard (27)

Patients were screened to identify those admitted with signs/symptoms of acute respiratory disease. Only patients with consolidation (radiologically proven respiratory infection) or opacities consistent with COVID19 pneumonitis on admission radiology were included. Pneumonia was defined as new radiological shadowing with no alternate explanation in the opinion of a consultant radiologist^5^. Patients with hospital-acquired pneumonia were excluded from this audit. COVID19 radiological change was defined as multiple bilateral peripheral opacities or other change in keeping with SARS-CoV-2 infection^7^.

Clinical data were collected from medical records. Patient eligibility for repeat radiology was determined separately by two authors: if disagreement occurred then the case was discussed. In both pneumonia and COVID19, patient follow-up was classified as under hospital team (where radiology had to be booked) or GP (which required the discharge summary indicate repeat radiology was required). Follow-up was assessed at 6- or 12-weeks for lobar pneumonia and bilateral COVID19 pneumonitis respectively.

### Statistical analysis

Categorical variables were presented as counts with percentages; continuous data as means and standard deviations (SD) if normally distributed and medians and interquartile range (IQR) if not normally distributed following testing for normality using the Shapiro-Wilk Test. Proportions were compared using Chi-squared testing; comparison of medians using Mann-Whitney U-Test, with P<0.05 considered significant.

## RESULTS

Overall, 596 adults were hospitalised with CAP and 444 with COVID19 (Figure 3). There were differences in the number of patients admitted with CAP and COVID19 across the three audit periods (Figure 3). Patient demographics are listed in Table 1. Adults hospitalised with CAP were older, more likely to reside in a long-term care facility and had higher frailty scores than those admitted with COVID19. Additionally, the burden of pre-existing medical conditions was higher in patients hospitalised with CAP, reflected by the higher CCI score for these patients. Importantly, patients hospitalised with CAP had higher rates of existing malignancy in comparison to those with COVID19.

**Figure 3:**
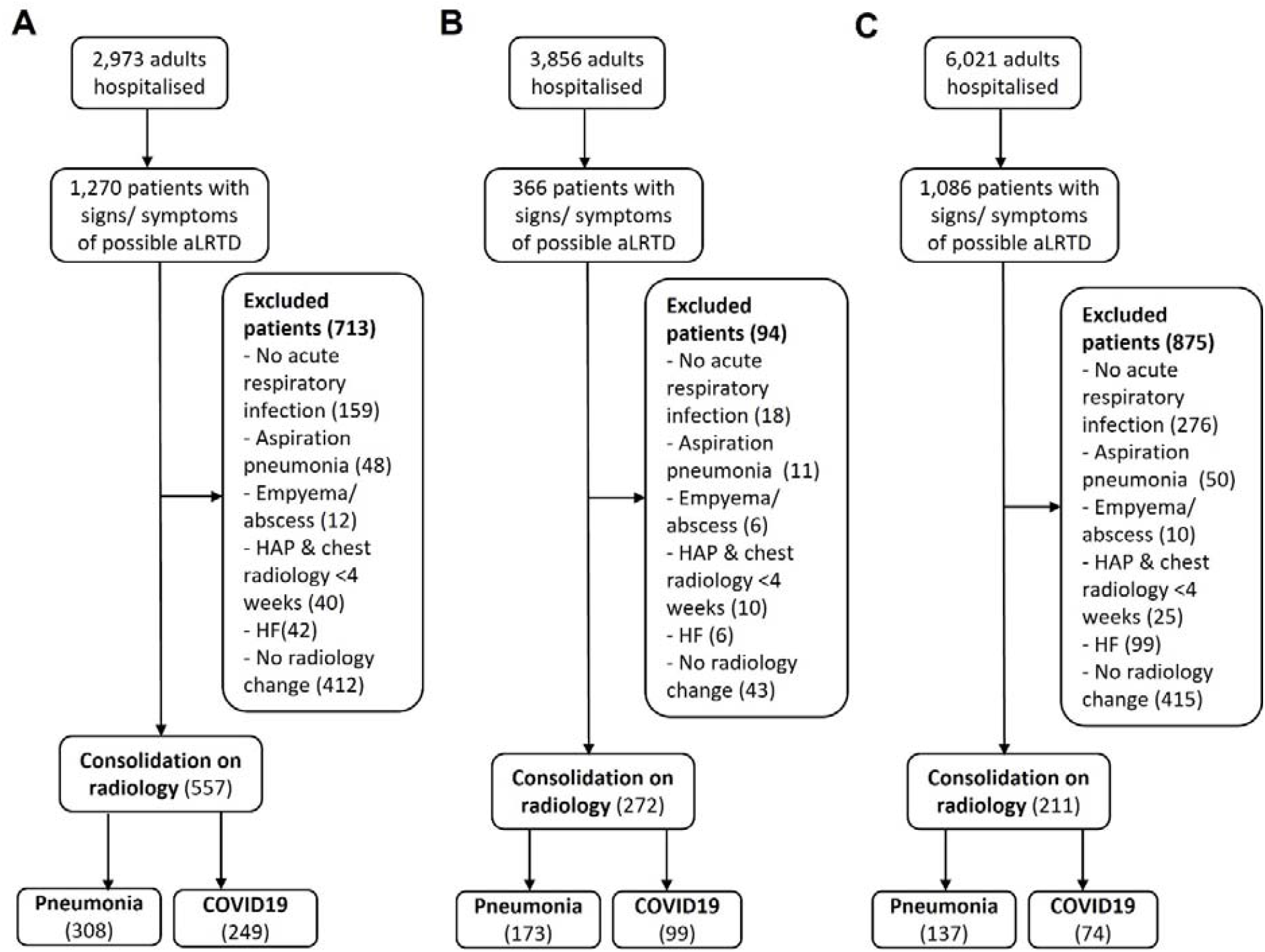
Study Flow Diagram. In this audit of adults hospitalised at North Bristol NHS Trust, there were (A) 557 adults in Aug-Dec 2020, (B) 272 adults in Jun-July 2021 and (C) 211 adults in Dec 21-Jan 22 with confirmed lobar pneumonia or pneumonitis on their initial chest radiology. aLRTD, acute lower respiratory tract disease; HAP, hospital acquired pneumonia; HF, heart failure

**Table 1:** Characteristics of Patients Hospitalised with Radiologically Proven Respiratory Infection.

| Characteristic | Cohort One (Aug-Dec 2020) |  |  | Cohort Two (June-Aug 2021) |  |  | Cohort Three (Dec-Jan 2022) |  |  |
| --- | --- | --- | --- | --- | --- | --- | --- | --- | --- |
|  | Pneumonia<br>N =308 | COVID-19<br>N =249 | Total Cohort<br>N =557 | Pneumonia<br>N = 173 | COVID-19<br>N = 99 | Total Cohort<br>N =272 | Pneumonia<br>N =137 | COVID-19<br>N = 74 | Total Cohort<br>N = 211 |
| Age (years) - median (IQR) | 80.0 (68.0–87.4) | 62.8 (54.2–76.9) | 73.7 (59.4–83.9) | 75.6 (61.8–85.7) | 48.5 (35.0–59.6) | 67.1 (48.5–81.7) | 75.8 (60.9–85.8) | 60.4 (53.5–78.2) | 68.9 (57.1–82.3) |
| Male Gender – N (%) | 159 (51.6) | 156 (62.7) | 315 (56.6) | 92 (53.2) | 62 (62.6) | 154 (56.6) | 69 (50.4) | 40 (54.1) | 109 (51.7) |
| Care home resident – N (%) | 48 (15.6) | 11 (4.4) | 59 (10.6) | 10 (5.8) | 0 (0) | 10 (3.7) | 22 (16.1) | 10 (13.5) | 32 (15.2) |
| **Smoking – N (%)** |  |  |  |  |  |  |  |  |  |
| Current | 43 (14.0) | 8 (3.2) | 51 (9.2) | 15 (8.7) | 5 (5.1) | 20 (7.4) | 8 (5.8) | 8 (10.8) | 16 (7.6) |
| Ex-smokers | 130 (42.2) | 113 (45.4) | 243 (43.6) | 94 (54.3) | 36 (36.4) | 130 (47.8) | 71 (51.8) | 26 (35.1) | 97 (46.0) |
| Unknown | 62 (20.1) | 32 (12.9) | 94 (16.9) | 13 (7.5) | 14 (14.1) | 27 (9.9) | 15 (10.9) | 10 (13.5) | 25 (11.8) |
| **Comorbidity Scores – N (%)** |  |  |  |  |  |  |  |  |  |
| Rockwood Frailty 0-4 | 120 (39.0%) | 203 (81.5%) | 323 (58.0%) | 103 (59.5) | 89 (89.9) | 192 (70.5) | 66 (48.2) | 55 (74.3) | 121 (57.3) |
| Rockwood Frailty 5-9 | 188 (61.0%) | 46 (18.5%) | 234 (42.0%) | 70 (40.5) | 10 (10.1) | 80 (29.4) | 71 (51.8) | 19 (25.6) | 90 (42.7) |
| CCI - median (IQR) | 5 (4 - 7) | 3 (1 - 4) | 4 (2 - 6) | 4 (2 - 6) | 1 (0 - 3) | 3 (1-5) | 4 (2 - 6) | 4 (2 - 6) | 4 (2 - 6) |
| **Respiratory – N (%)** |  |  |  |  |  |  |  |  |  |
| Any | 140 (54.5) | 68 (27.3) | 208 (37.3) | 81 (46.8) | 28 (28.3) | 109 (40.1) | 61 (44.5) | 25 (36.5) | 86 (61.1) |
| COPD | 95 (30.8) | 19 (7.6) | 114 (20.5) | 45 (26.0) | <5 (<5.1) | 49 (18.0) | 34 (24.8) | 13 (17.6) | 47 (22.2) |
| Asthma | 35 (11.4) | 39 (15.7) | 74 (13.3) | 34 (19.7) | 22 (22.2) | 56 (20.6) | 26 (19.0) | 9 (12.2) | 35 (16.6) |
| Other * | 35 (11.4) | 16 (6.4) | 51 (9.2) | 14 (8.1) | <5 (<5.1) | 16 (5.9) | 12 (8.8) | 5 (6.8) | 17 (8.1) |
| **Cardiovascular – N (%)** |  |  |  |  |  |  |  |  |  |
| CCF | 54 (17.5) | 16 (6.4) | 70 (12.6) | 21 (12.1) | <5 (<5.1) | 24 (8.8) | 23 (16.8) | 9 (12.2) | 31 (15.0) |
| Ischaemic Heart Disease | 62 (20.1) | 32 (12.9) | 94 (16.9) | 24 (13.9) | <5 (<5.1) | 28 (10.3) | 22 (16.1) | 9 (12.2) | 21 (10.0) |
| **Neurological – N (%)** |  |  |  |  |  |  |  |  |  |
| CVA/TIA | 39 (12.7) | 18 (7.2) | 57 (10.2) | 21 (12.1) | 0 (0) | 21 (7.7) | 14 (10.2) | 5 (6.8) | 19 (9.0) |
| Dementia | 40 (13.0) | 7 (2.8) | 47 (8.4) | 12 (6.9) | 0 (0) | 12 (4.4) | 12 (8.8) | 6 (8.1) | 18 (8.5) |
| Cognitive impairment | 21 (6.8) | 7 (2.8) | 28 (5.0) | 11 (6.4) | 0 (0) | 11 (4.0) | 5 (3.6) | 5 (6.8) | 10 (4.7) |
| Other neurological condition ‡ | 20 (6.5) | 10 (4.0) | 30 (5.4) | 7 (4.0) | <5 (<5.1) | 8 (2.9) | 11 (8.0) | 7 (9.5) | 18 (8.5) |
| **Malignancy – N (%)** |  |  |  |  |  |  |  |  |  |
| None | 257 (83.4) | 233 (93.6) | 490 (88.0) | 151 (87.3) | 93 (93.9) | 244 (89.7) | 118 (86.1) | 60 (81.1) | 178 (84.4) |
| Solid Organ Cancer | 42 (13.6) | 10 (4.0) | 52 (9.4) | 21 (12.4) | 5 (5.1) | 26 (9.6) | 15 (10.9) | 8 (10.8) | 23 (10.9) |
| Haematological Malignancy | 12 (3.6) | 7 (2.9) | 19 (3.5) | <5 (<2.8) | <5 (<5.1) | <5 (<1.8) | 5 (3.6) | 6 (8.1) | 11 (5.2) |

Patients with COVID19 who were admitted from June-Aug 2021 were younger than those hospitalised in Aug-Dec 2020 and Dec 20–Jan 2021 (P=0.036 and P=0.032 respectively) (Table 1). Throughout the audit, a high proportion of patients were deemed eligible for follow-up radiology (75% CAP vs 81% COVID19 Aug-Dec 20; 75% vs 93% Jun-Aug 21; and 75% vs 95% Dec 21–Jan 22) (Table 2). Of patients eligible for radiological follow-up, 67% (559/831) had follow-up planned by discharge from hospital. In total, 61% (273/447) patients with CAP had follow-up planned by the time of hospital discharge; in comparison, 75% (286/384) of adults hospitalised with COVID19 had radiology follow-up planned on discharge. In the first two audit periods, fewer CAP patients had radiology follow-up planned compared to COVID19 patients (Table 2). Overall repeat radiological examination occurred in 54% (446/831) eligible patients and a total of 47% (393/831) underwent investigation within BTS guideline timeframe (Table 2). Overall, 49% (220/447) patients with CAP and 59% (226/384) of COVID19 disease patients underwent planned follow-up. A total of 45% (201/447) eligible adults hospitalised with CAP and 43% (192/447) patients with COVID19 disease patients received follow-up within BTS guideline timelines.

**Table 2:** Radiological follow-up of patients with pneumonia and COVID-19.

| Radiological Follow-Up | Cohort One (Aug-Dec 2020) |  |  | Cohort Two (June-Aug 2021) |  |  | Cohort Three (Dec-Jan 2022) |  |  |
| --- | --- | --- | --- | --- | --- | --- | --- | --- | --- |
|  | Pneumonia<br>N =308 | COVID-19<br>N =249 | *P*-value | Pneumonia<br>N =173 | COVID-19<br>N =99 | *P*-value | Pneumonia<br>N =115 | COVID-19<br>N = 96 | *P*-value |
| **Patients Eligible under BTS Guidelines<sup>†</sup> - N (%)** |  |  |  |  |  |  |  |  |  |
| Not eligible | 77/308 (25) | 48/249 (20) | - | 43/173 (25) | 7/99 (7) | - | 29/115 (25) | 5/96 (5) | - |
| Eligible for follow-up | 231/308 (75) | 201/249 (81) | - | 130/173 (75) | 92/99 (93) | - | 86/115 (75) | 91/96 (95) | - |
| **Plan on Discharge - N (%)** |  |  |  |  |  |  |  |  |  |
| No follow-up booked | 106/231 (46) | 63/201 (31) | 0.0001 | 49/130 (38) | 20/92 (22) | 0.0116 | 17/86 (20) | 13/91 (14) | 0.2887 |
| Self-discharge before follow-up booked | 1/231(0) | 0/201 (0) | NS | 1/130 (1) | 0/92 (0) | NS | 0/86 (0) | 2/91 (2) | 0.1887 |
| Radiology plan implemented | 124/231 (54) | 138/201 (69) | 0.0015 | 80/130 (62) | 72/92 (78) | 0.0076 | 69/86 (80) | 76/91 (84) | 0.2980 |
| - Under hospital team | 72/124 (58) | 113/138 (82) | <0.0001 | 62/80 (78) | 67/72 (93) | 0.0097 | 58/69 (84) | 72/76 (95) | 0.1028 |
| - Under GP | 43/124(35) | 20/138 (14) | 0.0002 | 11/80 (14) | 5/72 (7) | NS | 6/69 (9) | 4/76 (5) | 0.5806 |
| - Documented plan not to repeat radiology | 9/124 (8) | 5/138(4) | NS | 7/80 (9) | 0/72 (0) | NS | 5/69 (7) | 0/76 (0) | 0.0493 |
| **Follow-up Occurrence - N (%)** |  |  |  |  |  |  |  |  |  |
| Total patients undergoing repeat radiology | 97/231 (42) | 118/201 (59) | 0.0004 | 64/130 (49) | 43/92 (47) | NS | 59/86 (69) | 65/91 (71) | 0.4122 |
| Follow-up occurred within timeframe | 87/231 (38) | 104/201 (52) | 0.0017 | 60/130 (46) | 25/92 (27) | 0.0042 | 54/86 (63) | 63/91 (69) | 0.2256 |
| - Under hospital team | 58/72 (81) | 93/113 (82) | NS | 51/62 (82) | 20/67 (30) | <0.0001 | 52/58 (90) | 61/72 (85) | 0.3047 |
| - Under GP | 29/43 (67) | 11/20 (55) | NS | 9/11 (82) | 5/5 (100) | NS | 2/6 (33) | 2/4 (50) | 1.0000 |

Both planning and occurrence of follow-up for adults with CAP increased between the audit period 1 and 2, alongside improvement of follow-up occurrence within BTS guideline timeframe. Planning of follow-up increased in patients hospitalised with COVID19, there was a decrease in occurrence of repeated radiological investigation from 59% to 47% of eligible individuals occurred. Of those with follow-up planned, most (80%) had this planned under the hospital team. Notably, 30% COVID19 patients admitted in June-Aug 2021 underwent follow-up per guidelines when arranged under the hospital team; in all other patient groups approximately 80% patients with hospital follow-up arranged underwent such follow-up. Patients with GP led follow-up tended to be less likely to receive follow-up within guideline criteria (P=0.03); although, there are small numbers in this audit group (Table 2).

In cohort 3, which was audited following the implementation of a policy which required radiological follow-up to occur under the discharging clinical team, there was noticeable increase in the proportion of follow-up for patients with CAP and COVID19 (69% and 71% versus 49% and 47% respectively in previous audit window). Whilst some follow-up still occurred led by the GP, this was lower than in other periods (Table 2).

## DISCUSSION

This single-centre audit found that initially only 49% of eligible patients under current BTS guidelines underwent such imaging. Appropriate patient follow-up is important to detect early and potentially treatable respiratory disease with 2.5% of chest radiographs repeated <90 days of pneumonia diagnosis result in lung cancer diagnosis, with cancer diagnostics predicted to increase if radiographs were only repeated in patients >50 years^4^. There is sparse literature concerning the occurrence rate of radiological follow-up after a diagnosis of pneumonia. A previous local audit showed 51% (55/107) patients underwent radiological follow-up within 6-weeks of presentation^10^. A separate audit found 43% patients ≥50y received appropriate advice concerning follow-up, with only 15% undergoing repeat imaging^11^. The level of adherence to BTS guidelines following identification of consolidative change is concerning: BTS CAP guidelines^5^ are well established, with repeated imaging to prevent delay in diagnosing lung cancer following demonstration of consolidation. Lung cancer remains the second most common cancer and accounts for 21% of all cancer deaths^12^ with 70% of lung cancer diagnosed when it has progressed to Stage 3 or 4^13^. Delay in diagnosis of lung cancer leading to more extensive malignancy at the time of diagnosis results in fewer curative treatment options and is associated with poor outcomes^14^. As such BTS guideline adherence should be high to permit early diagnosis and hopefully reduce lung cancer mortality. Further to this, we found 16% of booked follow-up did not take place and this needs to be addressed.

We found improvement in radiological follow-up rates both overall and within BTS timelines following implementation of a change in responsibility of radiology follow up booking at the study hospital. There were also some changes in guideline adherence between the first two audit periods, with planned follow-up for both COVID19 pneumonitis and CAP increasing between the audit periods. This may be attributable to the time periods which were audited, as UK medical rotations occur in August^15^ and therefore the second audit period occurred when doctors were more familiar with the hospital, guidelines or protocols and have received more training. As there are differences in the patient cohorts between the two audit periods, it is possible that the patient groups have different treatment preferences, and this affected patient attendance at follow-up appointments. For example, some patient groups may have higher rates of non-attendance at appointments^16,17^, or patients may have chosen to avoid hospital for repeated radiology testing as they were concerned about acquiring COVID19^18,19^.

This audit demonstrates that radiological follow-up was more likely to occur if arranged under the hospital team rather than under the GP. This may be because the hospital team directly book the radiology test at the same time as writing the discharge summary; whereas asking the patient’s GP to request follow-up radiology requires that GP undertake additional action to book the investigation. Additionally, it is possible that patients assign different significance to investigations depending on which clinician instigates the test, how the test and appointment are communicated to patients, or other factors such as perceived healthcare and NHS pressures may affect attendance at follow-up radiology appointments^19,20^. We highlight that only a small number of patients received follow-up under GP care; however, these results align with concerns about patient safety when hospital physicians request GPs to follow-up on tests requested during hospitalisation^21^. Significant improvement occurred in guideline adherence occurred when GPs were not asked to undertake this task.

In the first two audit periods, we also found that fewer CAP patients (45%, 161/361) underwent repeat chest radiology in comparison to those admitted with COVID19 (55%, 161/293), demonstrating difference in adherence between the two guidelines within the same hospital. Additionally, the hospital team planned follow-up under the GP more frequently for adults hospitalised with CAP than for those admitted with COVID19. The change implemented between audit period 2 and 3 not only increased total adherence but reduced the difference in follow-up rates between CAP and COVID19 patients. This audit did not investigate reasons for difference in CAP and COVID19 follow-up, but the time-period for COVID19 follow-up is longer than that for CAP^7^, and therefore it is possible pressure on healthcare had more effect on COVID19 follow-up. The increased prevalence of SARS-CoV-2 infection coupled with the more recent BTS guidelines^7^, in the context of frequent advances in knowledge and COVID19 disease treatment guidelines, may mean that physicians were more familiar with these guidelines or developed strategies to improve implementation of care pathways for patients with COVID19 disease, thus increasing adherence. It is also possible that the difference in follow-up planning may be attributable to physician and patient treatment preferences for frailer patients, adults with considerable pre-existing medical disease burden or those with malignancy. The known difference in circulating and dominant SARS-CoV-2 variants may have affected either patient or physician treatment preference.

The follow-up rate found in our audit during this time was higher than that found in a study conducted at a UK district general hospital, which reported 58% of adults with COVID19 underwent radiological follow-up^22^. As North Bristol NHS Trust has a dedicated specialist pulmonary fibrosis centre, radiological follow-up to detect early change following COVID19 may be higher than in non-specialist units, in line with studies demonstrating that specialist units may offer improved patient care^23,24^.

This audit has many strengths. Firstly, it was conducted at a University hospital with specialist respiratory services and recruited many patients with CAP and COVID19. We repeated the audit over two time periods at the same hospital, showing similar results within the same healthcare setting. The first two time periods occurred within the same training/rotation year; therefore, this audit was repeated with the same cohort of junior doctors in post and any change in practice occurred within the same group of physicians.

There are also some limitations: as this is a single-centre audit, results may not be generalisable. By necessity we audited time periods when the NHS was under considerable pressure, and therefore rates of appropriate follow-up may improve when services are not under such strain. We were unable to fully determine reasons for non-adherence to BTS guidelines in this audit, including why radiological follow-up was not arranged, or why patients did not receive follow-up, which may have been due to patient preference. By necessity, this audit took place in time periods during which different SARS-CoV-2 variants were circulating and/or dominant.

## CONCLUSION

In conclusion, these data suggest BTS guideline adherence regarding radiological follow-up may be suboptimal, with room for improvement and consequently improved patient outcomes. We found that adherence to BTS guidelines improved, once it was established that this follow-up should be arranged by the inpatient clinical team. This audit should be undertaken at additional hospitals, and potentially nationally, to ensure appropriate BTS guidelines adherence thereby avoiding delay in diagnosing underlying malignancy or chronic lung disease. Furthermore, additional reasons for non-adherence to guidelines should be determined as this will allow these issues to be addressed and the treatment of patients improved.

## Data Availability

No additional data available.

## Notes

**Declaration of Interests:** CH is principal investigator of the Avon CAP study which is an investigator-led University of Bristol study funded by Pfizer and has previously received support from the NIHR in an academic clinical fellowship. The other authors have no relevant conflicts of interest to declare.

### Competing Interest Statement

CH is principal investigator of the Avon CAP study which is an investigator-led University of Bristol study funded by Pfizer and has previously received support from the NIHR in an academic clinical fellowship. The other authors have no relevant conflicts of interest to declare.

### Author Declarations

North Bristol NHS Trust Research and Audit Ethics Committee, CA92571

### Summary of Updates

Updated to reflect some changes following review

